# Identification of pleiotropic genes and biological mechanisms underlying the shared genetic component between asthma, neuropsychiatric disorders, and substance use behaviors

**DOI:** 10.64898/2026.10.16.26364574

**Authors:** Haibo Huang, Raphaël Vernet, Lucie Troubat, Christophe Linhard, Lucas Chataigner, Yuka Suzuki, Stephan Fischer, Hugues Aschard, Florence Demenais, Emmanuelle Bouzigon, Hanna Julienne

**Affiliations:** Université Paris Cité, INSERM, HealthFex, Genomic Epidemiology of Multifactorial Diseases Group, Paris, France; Institut Pasteur, Université Paris Cité, Department of Computational Biology, Paris, France

## Abstract

Asthma patients suffer from neuropsychiatric disorders (NPDs) more often than expected and NPD patients have a higher prevalence of smoking, which can lead to asthma. To investigate the genetic mechanisms underlying this complex relationship, we performed an integrative analysis of GWAS summary statistics from four asthma subtypes, 12 NPDs, and six substance use behaviors (SUBs). Asthma shares an important genetic component with post-traumatic stress disorder (PTSD), major depressive disorder, and attention-deficit/hyperactivity disorder (ADHD) that cannot be fully accounted for by smoking. Multi-trait GWAS identified 354 independent variants mapping to 178 pleiotropic genes capturing the core of the shared genetics of asthma, NPD, and SUB. Single-cell transcriptomic analysis across blood, brain, and lung tissues revealed pleiotropic gene overexpression in 57 cell types and three candidate mechanisms linking asthma and NPDs. First, immune-related pleiotropic genes are expressed in microglia, blood and lung immune cells, suggesting shared immune dysfunction. Second, the pleiotropic genes *EP300*, *RERE*, and *MAML3* regulate NOTCH signaling and are expressed in neurons, microglia, and lymphoid progenitors. Third, pleiotropic genes expressed in ciliated cells of both lung and brain implicate ciliary dysfunction. These findings may help identify new treatment strategies for patients with co-occurring asthma and neuropsychiatric disorders.

## INTRODUCTION

Asthma is the most prevalent inflammatory lung disease, affecting more than 262 million people worldwide^1^. Resulting from multiple genetic and environmental factors and their interplay, asthma is a heterogeneous disease with variable clinical and biological presentations over the life span^2^, which is partly due to genetic heterogeneity^3^. It can be divided into subtypes according to various clinical and biological features, including the age at asthma onset (e.g. childhood or adulthood), inflammatory profile (e.g. eosinophilic or neutrophilic), disease severity, and comorbidities^2^.

Epidemiological studies evidence that asthma and neuropsychiatric disorders often co-occur^4^. Patients with asthma have higher prevalence of mental disorders (e.g. Odds ratio equal to 1.64-1.67, 1.45-1.95, 1.34-1.53 and 5.38 for major depression, anxiety, ADHD and PTSD respectively^4–7^). Similarly, people with mental health conditions are at an increased risk of developing asthma (e.g. hazard ratio equal to 1.65, 1.68, 1.72 and 1.94 for prior schizophrenia, mood disorder, behavioral and personality disorder respectively)^8^. Some of these associations predominantly emerge in childhood while others are more common in later life^8^. Recent studies provide evidence of bidirectional associations between asthma and numerous mental disorders, suggesting possible shared etiological or pathophysiological processes^8^. Indeed, a cross-talk could occur between the central nervous system and the lung through several pathways, including neuroanatomical, endocrine, immune, metabolite, microorganism or gas pathways, as illustrated by the complex lung-brain axis^9,10^. Beyond epidemiological associations, increasing evidence supports shared genetic components between asthma and neuropsychiatric conditions^11,12^. Importantly, part of these associations may be influenced by substance use behaviors, such as smoking, which is more frequent in neuropsychiatric patients^13–15^ and directly contributes to lung damage and increases the risk of asthma^16^.

To our knowledge, the potential contributing role of smoking in the shared genetic etiology between asthma and neuropsychiatric disorders has never been investigated. In addition, the shared genetic architecture between asthma and certain neuropsychiatric disorders, such as obsessive-compulsive disorder, remains unexplored. Understanding of the shared genetic architecture between asthma and neuropsychiatric disorders, as well as the role/implication of substance use behaviors in this relationship, has important clinical implications: (1) to determine whether one type of disease may represent a risk of another, (2) to unveil shared molecular insights of comorbidities^17^. Moreover, by revealing shared disease mechanisms, it can identify novel drug targets or repurpose existing drugs based on pleiotropic genes, as well as facilitating the identification of valuable targets for multiple disease outcomes^10,17,18^. Finally, it will allow the identification of patient subsets with potential implications for personalized medicine.

Here, we performed an integrative analysis of large GWAS summary statistics for four asthma subtypes (ever asthma, adult-onset, childhood-onset and moderate-to-severe asthma), 12 neuropsychiatric disorders (NPDs), and six substance use behaviors (SUBs), in order to characterize the shared genetic components across these traits at the genome-wide scale. These analyses included a wide range of complementary multivariate descriptors, including genome-wide and local genetic correlations, multi-trait pairwise GWAS analysis between asthma subtypes and NPDs/SUBs, colocalization analysis, functional annotations and single-cell transcriptomic mapping in blood, brain, and lung tissues.

## Results

### Overview of the study

This study had two main objectives. First, we aimed to describe the genetic relationships between asthma subtypes and neuropsychiatric disorders (NPDs) while taking into account the potential mediating effect of substance use behaviors (SUBs), notably smoking behaviors. Second, we aimed to uncover the genes, cell types and pathways underlying these shared genetic components. To achieve these objectives, we performed an integrative analysis comprising four main steps (Fig. 1, Supplementary Fig. 1) using European-ancestry GWAS summary statistics (Supplementary Data 1) of four asthma subtypes (ever asthma, adult-onset asthma, childhood-onset asthma, and moderate-to-severe asthma), 12 neuropsychiatric disorders, and six substance use behaviors. NPDs included four internalizing disorders (anxiety disorder, panic disorder, PTSD, and major depressive disorder), four compulsive disorders (obsessive compulsive symptom, obsessive compulsive disorder, eating disorder, and tourette syndrome), two neurodevelopmental conditions (autism spectrum disorder and ADHD), schizophrenia, and bipolar disorder. SUBs included smoking and drinking behaviors, and cannabis use disorder (see Supplementary Note 1 for detailed definition).

**Fig. 1.**
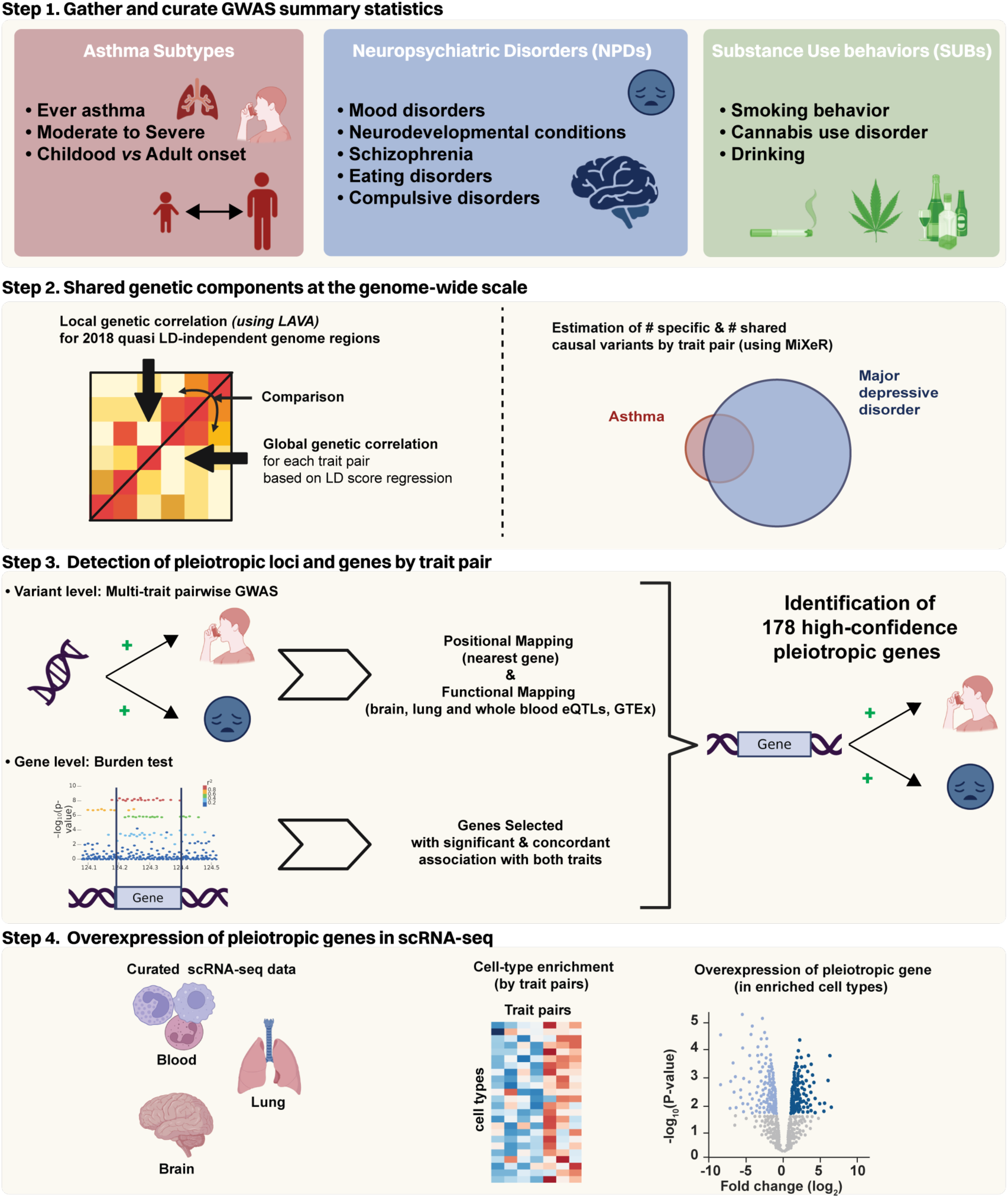
| Study overview. We analyzed the shared genetic etiology between asthma and neuropsychiatric disorders through four main steps. 1) We curated 24 GWAS summary statistics covering asthma subtypes, neuropsychiatric disorders (NPDs), and substance use behaviors (SUBs). All GWAS were harmonized and imputed using the JASS pipeline. 2) We assessed the shared genetic etiology at the genome-wide scale using complementary tools: the LD score regression-based genetic correlations, local genetic correlations as implemented in LAVA, and the number of shared causal variants estimated by bivariate MiXeR. 3) For each trait pair, we detected pleiotropic variants associated with increased risk in both traits. Pleiotropic variants were mapped to genes based on position, and eQTL data. We also performed a gene-based burden test to identify additional pleiotropic genes. 4) We performed the functional annotation of the 178 high confidence pleiotropic genes by analyzing their expression in single cell RNA-seq data across three relevant tissues (brain, lung, and blood). We computed cell-type enrichments for each gene set associated with a trait pair and investigated the over-expression of pleiotropic genes in enriched cell types.

We first characterized the shared genetic components across these traits using several complementary methods (Fig. 1, step 2): LD-score regression-based genetic correlations over the entire genome^19^, local genetic correlations as implemented in LAVA^20^, and bivariate MiXeR^21^, a Bayesian approach estimating the number of shared causal variants between two traits. We then (Fig. 1, step 3) detected pleiotropic variants through bivariate GWAS, and pleiotropic genes through a combination of variant-to-gene assignment and gene-based burden tests. Finally (Fig. 1, step 4), we identified candidate cell types and pathways underlying the shared etiology between asthma subtypes, NPDs, and SUBs through the integration and analysis of single cell RNA-seq datasets in three tissues: blood^22^, lung^23^, and brain^24^.

### At the genome-wide scale, the overlap between asthma subtypes and neuropsychiatric disorders or substance use behaviors is stronger for adult-onset asthma and moderate-to-severe asthma than for childhood-onset asthma

We observed substantial global genetic correlations between three asthma subtypes (ever asthma, adult-onset asthma, and moderate-to-severe asthma) and several neuropsychiatric disorders (Fig. 2a, Supplementary Data 2**-**4, see Methods): major depressive disorder (0.26 ≤ *r_g_* ≤ 0.37, *P*_FDR_ ≤ 3.83x10^-13^), PTSD (0.28 ≤ *r_g_* ≤ 0.40, *P*_FDR_ ≤ 6.91x10^-11^), and ADHD (0.26 ≤ *r_g_* ≤ 0.36, *P*_FDR_ ≤ 3.38x10^-8^). While less significant, there are also substantial genetic correlations between anxiety disorders and the same asthma subtypes (0.35 ≤ *r_g_* ≤ 0.48, *P*_FDR_ ≤ 0.04). Genetic correlations between asthma subtypes, schizophrenia, bipolar disorder, autism spectrum disorder, tourette syndrome, and panic disorder are more moderate (*r_g_* ≤ 0.2). In contrast, childhood-onset asthma shows significant correlations only with major depressive disorder and PTSD, and to a lesser degree (*r_g_* ≤ 0.06, 0.03 ≤ *P*_FDR_ ≤ 0.04), consistent with previous studies^12,25^. Eating disorder, obsessive compulsive symptom, and obsessive-compulsive disorder are not correlated with any asthma subtypes. Asthma subtypes also display genetic correlation with some SUBs. Ever asthma and adult-onset asthma correlate with smoking initiation, age-at-smoking initiation, and cigarettes smoked per day (-0.20 ≤ *r_g_* ≤ 0.28, 5.07x10^-23^ ≤ *P*_FDR_ ≤ 3.64x10^-2^). Moderate-to-severe asthma correlates with smoking initiation and cannabis use disorder (0.19 ≤ *r_g_* ≤ 0.28, 2.62 x10^-5^≤ *P*_FDR_ ≤ 2.89x10^-3^).

**Fig. 2.**
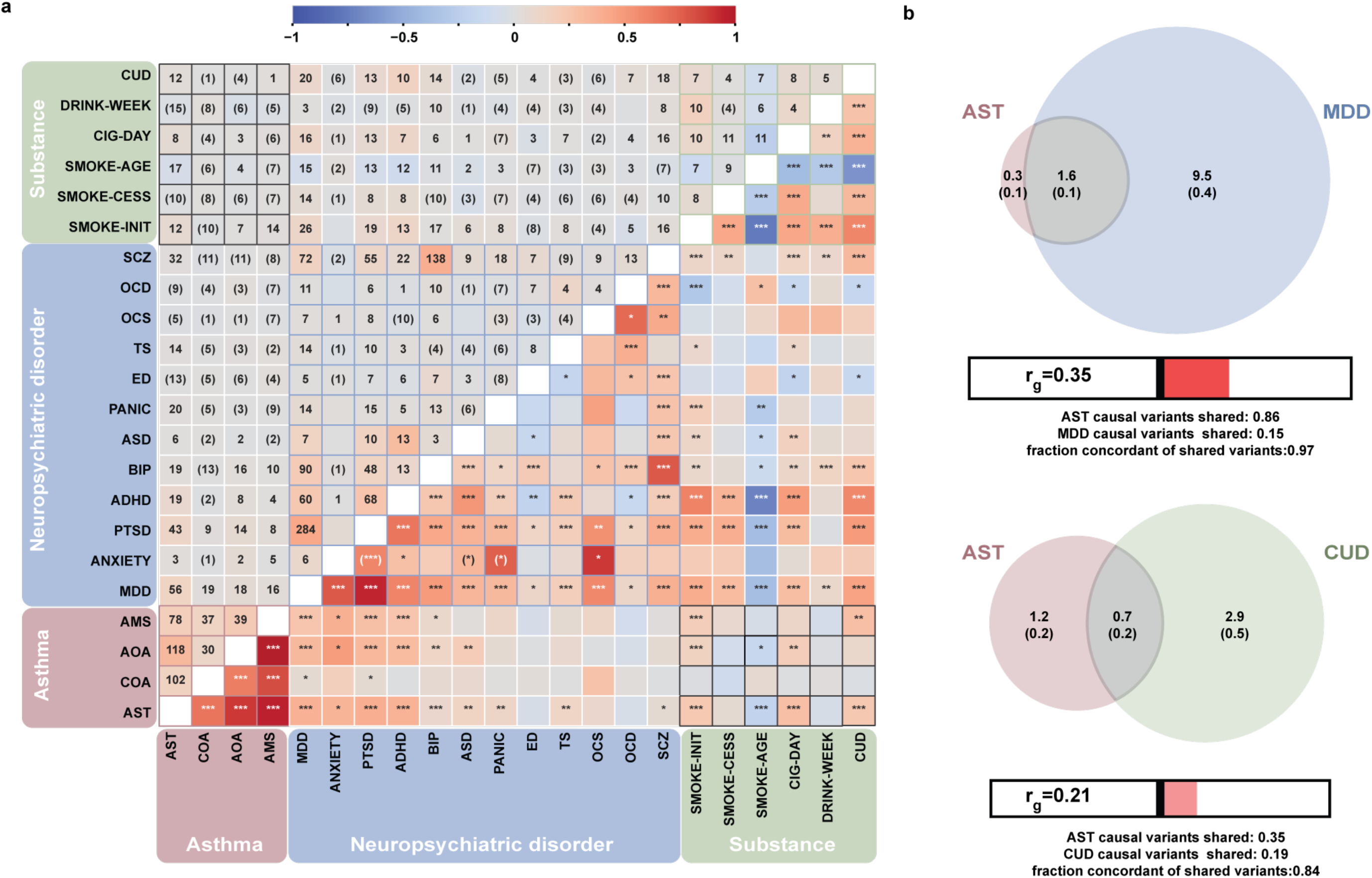
| Shared genetic components across trait pairs. **a)** Heatmap of global and local genetic correlations. Local genetic correlations are shown above the matrix diagonal (mean value across LD-independent region), while global genetic correlations are displayed below the diagonal. Significant global genetic correlations are denoted by asterisks (*: *P*_FDR_ < 0.05, **: *P*_FDR_ < 0.005, and ***: *P*_FDR_ < 0.0005). The number of regions with a significant local genetic correlation (*P*_FDR_ < 0.05) is annotated for each trait pair. Parentheses around region numbers indicate trait pair with non-significant global genetic correlations, while those around asterisks indicate trait pair with no significant local genetic correlations. **b, c**) Bivariate MiXeR estimates of genetic overlap between ever asthma and major depressive disorder (b) and ever asthma and cannabis use disorder (c). Venn diagrams display the estimated number of causal variants (in thousands; standard deviation in parenthesis) affecting both traits (grey), specific to ever asthma (pink), and specific to major depressive disorder (blue) or cannabis use disorder (green). Trait acronyms used in this figure: ever asthma: AST, adult-onset asthma: AOA, childhood-onset asthma: COA, and moderate-to-severe asthma: AMS, anxiety disorders: ANX, panic disorder: PANIC, post-traumatic stress disorder: PTSD, major depressive disorder: MDD, compulsive symptoms: OCS, obsessive compulsive disorder: OCD, eating disorder: ED, Tourette syndrome: TS, autism spectrum disorder: ASD, attention-deficit/hyperactivity disorder: ADHD, schizophrenia: SCZ, bipolar disorder: BIP, smoking initiation: SMOKE-INIT, smoking cessation: SMOKE-CESS, age-at-smoking initiation: SMOKE-AGE, number of cigarettes smoked per day: CIG-DAY, number of drinks consumed per week: DRINK-WEEK, and cannabis use disorder: CUD.

We estimated the shared and specific polygenic components for each trait pair with a sufficient sample size using MiXeR^21^ (see Methods). Overall, neuropsychiatric disorders are highly polygenic with an estimated number of causal variants above 7,000 (Supplementary Fig. 3). In contrast, asthma subtypes are less polygenic with less than 2,000 estimated causal variants for ever-asthma. Between 69% and 93% of ever asthma causal variants are shared with major depressive disorder, PTSD, ADHD, bipolar disorder, schizophrenia (Fig. 2b, Supplementary Fig. 3). However, because of their higher polygenicity, NPDs typically harbor a large specific polygenic component that does not impact asthma ranging from 80% to 85% of all their causal variants. The fraction of shared causal variants between asthma and NPDs with concordant effect sign was especially high for major depressive disorder, PTSD, and ADHD (above 95%) as compared to bipolar disorder, and schizophrenia (57% and 54%, respectively). Adult-onset asthma and moderate-to-severe asthma exhibit a comparable pattern of shared and trait-specific polygenic components with these neuropsychiatric disorders. In contrast, compared to other asthma subtypes, childhood-onset asthma shares a smaller fraction of causal variants with major depressive disorder, and ADHD (ranging from 31% to 51% versus > 69%), and a stronger overlap with obsessive-compulsive symptoms than the other asthma subtypes (63% *versus* < 28%).

The fraction of asthma subtype causal variants shared with substance use behaviors was higher for adult-onset asthma and moderate-to-severe asthma than for ever-asthma and childhood-onset asthma. For instance, adult-onset asthma and moderate-to-severe asthma share approximatively 70% of causal variants with cannabis use disorder, of which more than 72% show concordant effect sign (Supplementary Fig. 4), whereas ever asthma and childhood-onset asthma share less than 35% of causal variants with cannabis use disorder. The fraction of asthma subtypes (except childhood-onset asthma) causal variants shared with smoking initiation is also high; however, the broad definition of smoking initiation phenotype, encompassing smoking for a few months to a lifetime (Supplementary Note 1), complicates the interpretation of this trend. The overlap and genetic correlations between substance use behaviors and neuropsychiatric disorders are substantial (Fig. 2a, Supplementary Fig. 5, Supplementary Note 4), supporting the strong genetic connections between substance use behaviors and neuropsychiatric disorders. Thus, we might posit that the association between asthma and neuropsychiatric trait is mediated by substance use behaviors. However, since ever asthma shares 1639 causal variants with major depressive disorder and 671 causal variants with cannabis use disorder, more than half of causal variants of ever asthma are estimated to be shared with major depressive disorder but not with cannabis use disorder (Fig. 2b). This observation highlights that while substance use behaviors might contribute to the pleiotropy between asthma and neuropsychiatric disorders, they might not fully account for it.

### Local genetic correlations offer a different perspective on the patterns of correlations between asthma subtypes and other traits

Since global genetic correlations represent the average shared genetic effects across the genome, we further computed local genetic correlations for each trait pair across LD-independent genomic regions using LAVA^20^ (Fig. 2a, Supplementary Fig. 2, Supplementary Data 5, see Methods). We identified 666 significant (*P*_FDR_ <0.05) local genetic correlations between asthma subtypes and other traits. Of these, 245 (36.8%) are observed for trait pairs with a non-significant global genetic correlation (Supplementary Data 6). Notably, childhood-onset asthma shows a comparable number of significant local genetic correlations to adult-onset asthma across NPDs and SUBs: 11 vs. 11 with schizophrenia, 13 vs. 16 with bipolar disorder, 19 vs. 18 regions with major depressive disorder, and 10 vs. 7 with smoking initiation.

### Bivariate GWAS between asthma subtypes and neuropsychiatric disorders or substance use behaviors

To pinpoint further the loci shared between asthma subtypes and other traits, we conducted bivariate GWAS, testing association between each variant for every pair of phenotypes (asthma subtypes-NPDs and asthma subtypes-SUBs, 72 trait pairs) – mimicking the design of the local and global genetic correlation estimation and thus allowing for direct comparison – followed by colocalization analysis (see Methods). We focused on statistically significant variants after Bonferroni correction for this bivariate omnibus test (5x10^-8^ / 72) and with the same genetic effect sign for both traits (i.e., risk increasing or risk decreasing for both traits). We identified 354 significant associations corresponding to 270 unique lead variants (Figs. 3a and b, Supplementary Data 7), which we refer further as pleiotropic variants. Among these, 11 are novel associations (significant for the bivariate test but not for either univariate test, Supplementary Data 8, Supplementary Fig. 6). We analyzed the overlap between significant single-trait LD independent variants and pleiotropic variants. The observed trend is consistent with trends observed at the genome wide scale using MiXeR: the genetic overlap between ever asthma and neuropsychiatric disorders is larger than the genetic overlap between ever asthma and substance use behaviors (Supplementary Note 5).

**Fig. 3.**
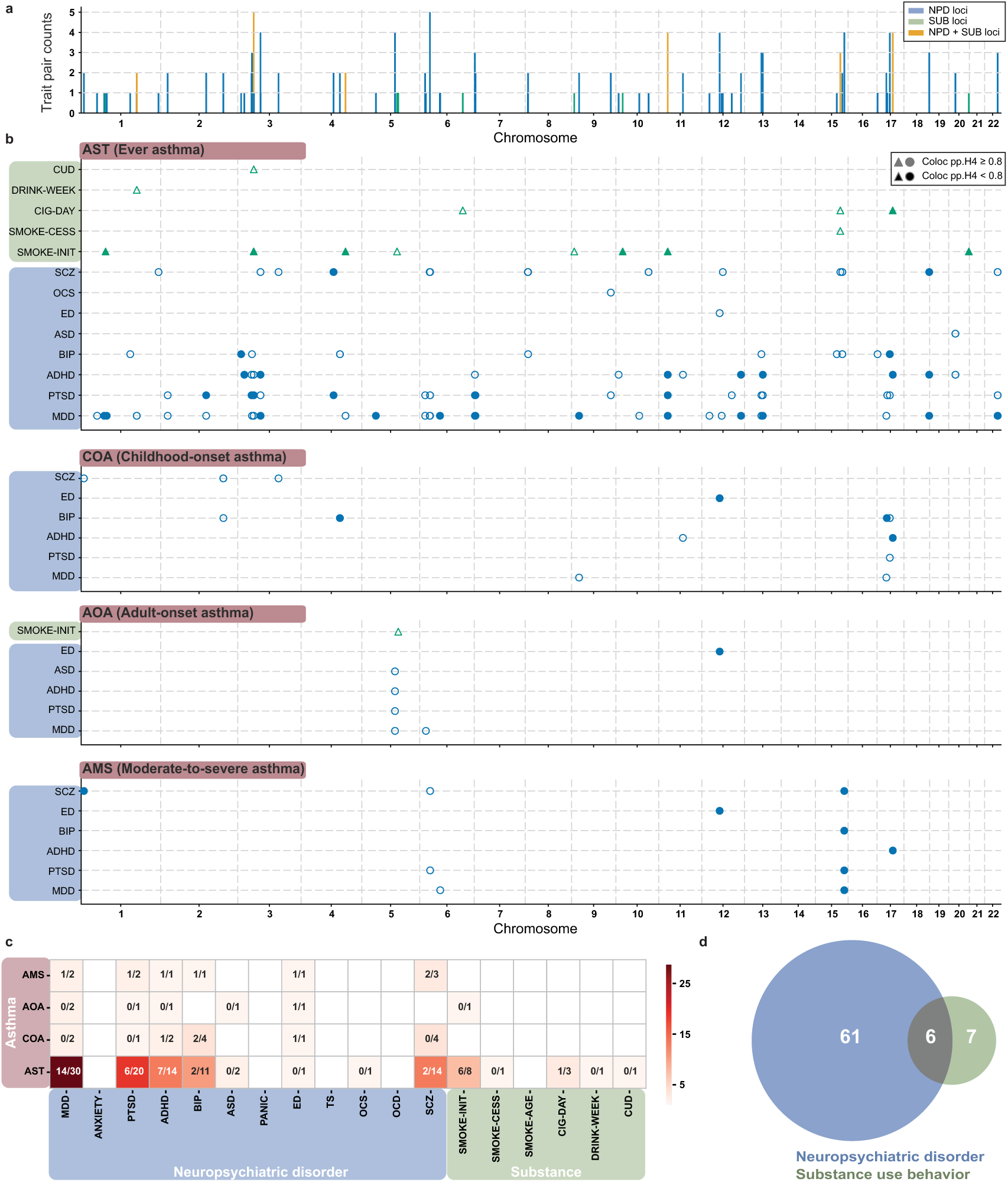
| Pleiotropic variants across trait pairs. **a)** Bar plot showing the number of trait pairs associated with each pleiotropic locus. Bar colors indicate the trait groups associated with the locus. **b)** Scatter plots showing the genomic position of lead pleiotropic variants identified across all trait pairs between asthma subtypes and NPDs (blue circles) or SUBs (green triangles). Panels, from top to bottom, represent ever asthma, childhood-onset asthma, adult-onset asthma, and moderate-to-severe asthma. Filled markers indicate variants that met the colocalization threshold (Coloc pp.H4 ≥ 0.8), whereas open markers indicate variants that did not. **c)** Heatmap showing the number of pleiotropic loci associated with each trait pair across the genome. In each cell, the annotation on the right is the total number of pleiotropic loci associated with each trait pair, while the annotation on the left is the number of pleiotropic loci that met colocalization threshold (i.e., at least one pleiotropic variant within that locus passed the colocalization test for the corresponding trait pair). The color scale represents the total number of pleiotropic loci, **d)** Venn diagram of the number of pleiotropic loci associated with asthma subtypes and NPDs (blue circle), and asthma subtypes and SUBs (green circle).

We merged lead variants located within 250 kb of one another into pleiotropic loci, yielding 74 loci in total (Fig. 3c, Supplementary Data 9). Ever asthma and major depressive disorder on one hand, and ever asthma and PTSD on the other hand, share the highest number of pleiotropic loci (30 and 20, respectively). We found 14 loci associated with increased risk of both schizophrenia and ever asthma, which confirm their shared genetic etiology despite a modest genetic correlation (*r_g_* =0.06). Similarly, the bivariate GWAS confirmed the existence of four pleiotropic loci raising the risk of childhood-onset asthma and bipolar disorder. Of the 74 pleiotropic loci, 67 are associated with asthma subtypes and neuropsychiatric disorders; 13 are associated with asthma subtypes and substance use behaviors; and only six are associated with all three groups (Fig. 3d). This further suggests a larger genetic overlap between asthma subtypes and neuropsychiatric disorders, than between asthma subtypes and substance use behaviors.

We also observed highly pleiotropic loci (Supplementary Fig. 8, Supplementary Data 9). Nine of these are significantly associated with more than four trait pairs (Table 1). The most pleiotropic locus located on Chr3-50Mb (Supplementary Fig. 12) lies within a dense gene region associated with ever asthma, three neuropsychiatric disorders (PTSD, major depressive disorder, ADHD) and two substance use behaviors (smoking initiation and cannabis use disorder). In this locus, several multi-trait lead variants met the colocalization threshold (Table 1) for ever asthma & smoking initiation (rs2352974, rs34654589, rs9843653, Supplementary Data 7), and ever asthma & PTSD (rs3774750, rs2073726). The locus Chr15-91Mb, is associated with moderate-to-severe asthma and four neuropsychiatric disorders (PTSD, major depressive disorder, bipolar disorder and schizophrenia) with a high colocalization probability across all trait pairs (pp.H4 > 0.97, Supplementary Data 7) and showing a peak signal in a region encompassing *FES* and *FURIN* genes (Supplementary Fig. 9). Whereas the univariate signal for moderate-to-severe asthma is not genome-wide significant, the bivariate GWAS signal is consistently significant across trait pairs. In addition, we noted that the bivariate lead variant (rs6224) being in strong LD with variants previously reported associated with asthma^26^ (D’=0.86, R²=0.46 with rs8029440) and neutrophil count^27^ (D’=1, R²=0.99, with rs8039305). The Chr3-71MB locus is associated with ever asthma, ADHD, major depressive disorder, schizophrenia, and PTSD (Supplementary Fig. 11) with high colocalization probabilities for ever-asthma, ADHD, and major depressive disorder (pp.H4 ≥ 0.92, Supplementary Data 7). The four multi-trait lead variants (rs13096070, rs13327339, rs7638354, rs62244862) in this locus lie in introns of the *FOXP1* gene.

**Table 1.**
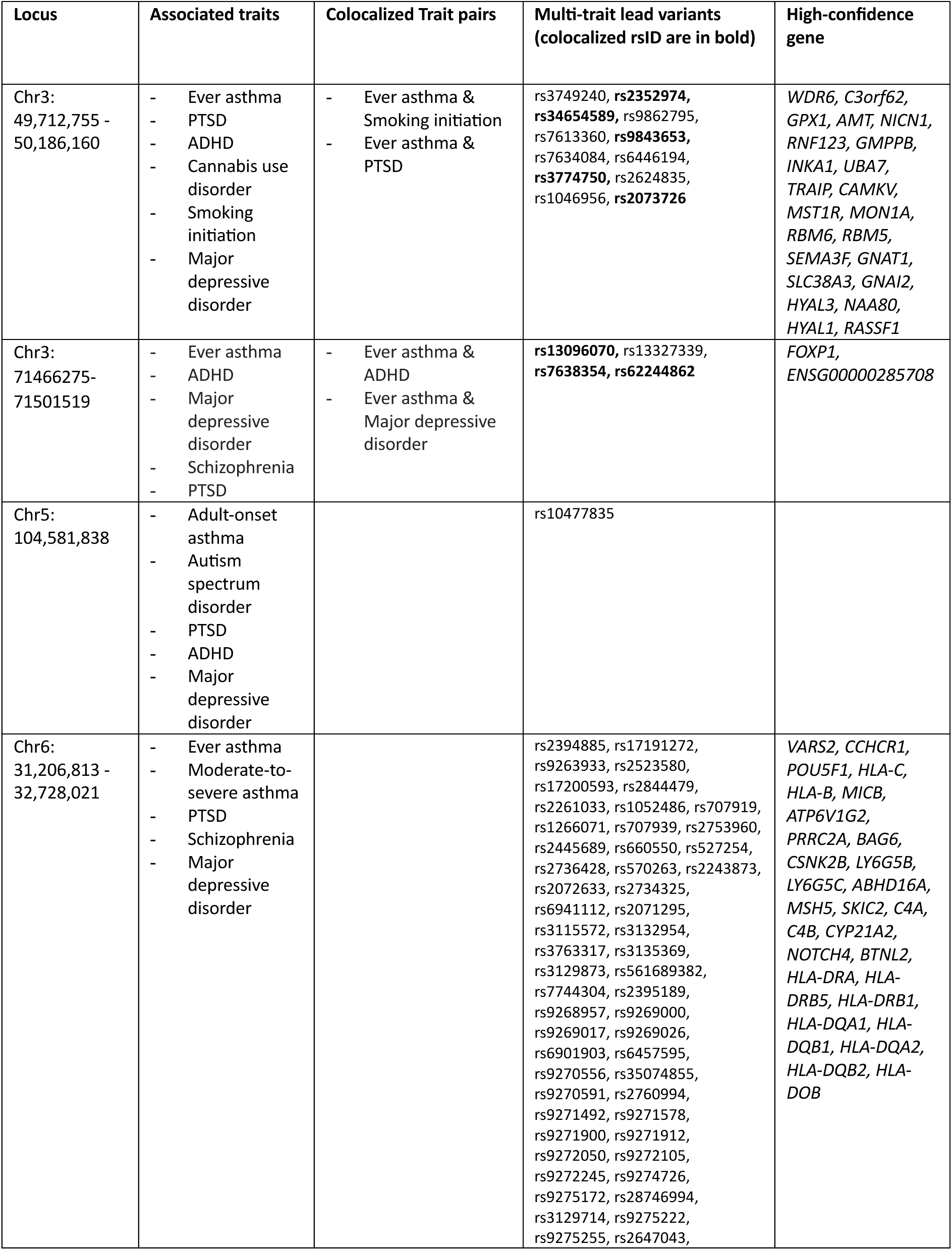
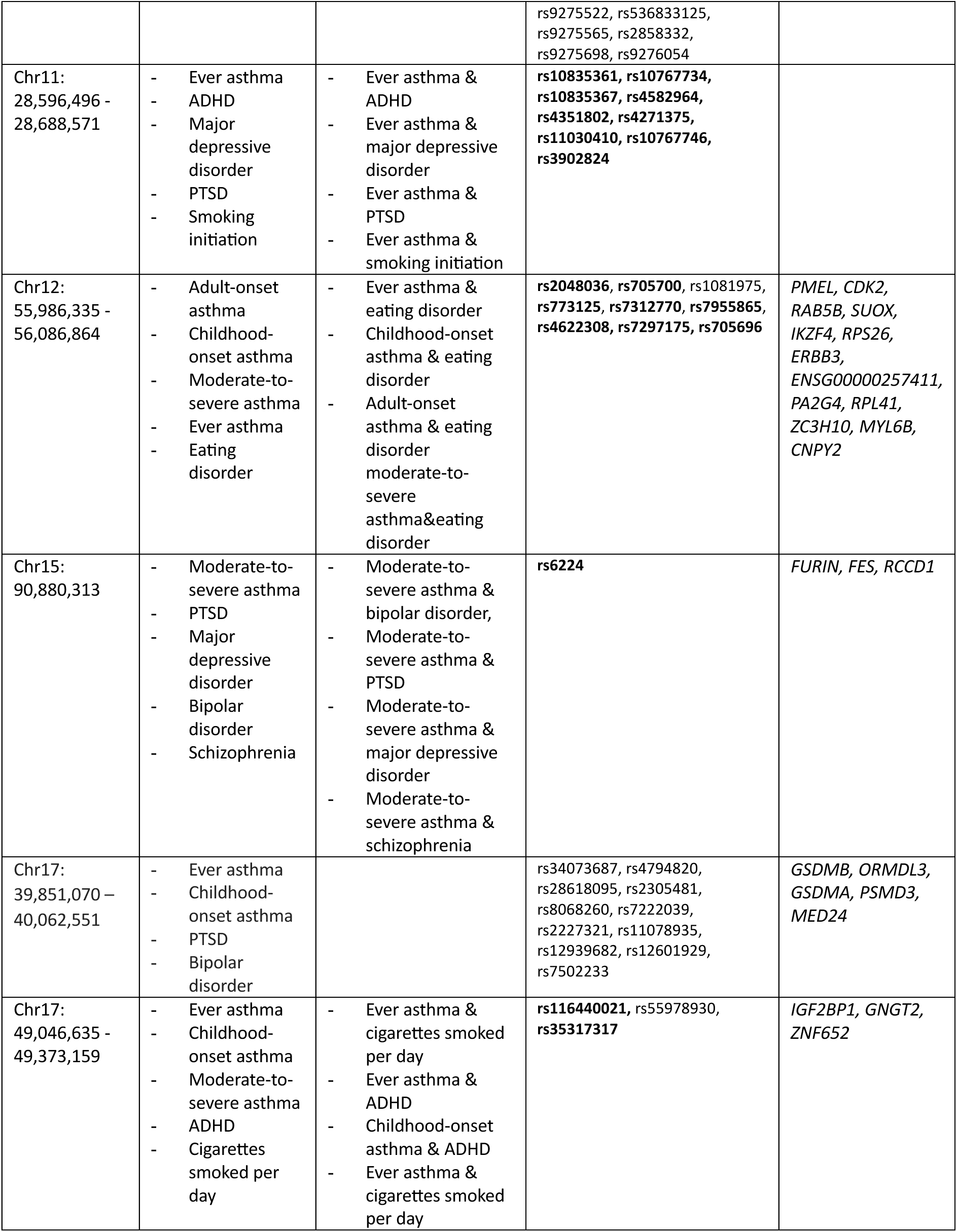
Nine pleiotropic loci associated with at least four traits. We considered a trait pair to be colocalized if at least one variant within the locus passed the colocalization test (pp.H4 ≥ 0.8). Similarly, we considered an rsID to be colocalized if it passed the colocalization test for at least one trait pair.

### Comprehensive functional analysis of pleiotropic variants

To understand the biological mechanisms impacted by the 270 pleiotropic variants, we conducted a comprehensive functional analysis including overlap with regulatory elements, gene mapping, single-cell analysis, and differential gene expression across cell types.

#### Annotation of pleiotropic variants

We first characterized the functional consequences of pleiotropic variants using the Ensembl Variant Effect Predictor (see Methods, Supplementary Fig. 13, Supplementary Data 10). Most pleiotropic variants are in gene introns (N=173) or nearby a gene (N=135, variants up to 5kb upstream or downstream of a gene) with a limited number lying in intergenic regions (N=31). Notably, three are missense variants, including rs1046956 (associated with ever asthma, ADHD, and smoking initiation) in the *SEMA3F* gene within the most pleiotropic locus we detected (Chr3-50Mb). In addition, we examined the RegulomeDb^28^ functional categories to which pleiotropic variants belong (Supplementary Data 7) and observed strong enrichments for the most functional categories (i.e., categories 1a to 1f, OR > 3.16, *P* < 0.02, Supplementary Fig. 13c) as compared to all variants scored in RegulomeDb. This implyies that our pleiotropic variants often overlap with previously reported eQTLs, transcription factor binding sites, and accessible chromatin marks.

#### Identification of pleiotropic genes

By using an integrative approach combining positional mapping, GTEx cis-eQTLs data in three relevant tissues (lung, brain, and blood), and a burden test^29^ (see Methods; Supplementary Fig. 1; and Supplementary Data 7, 11, 12), we identified a total of 315 pleiotropic genes, of which 178 are significant across multiple methods and, thus, are considered high confidence (see Methods, Supplementary Data 13). Twenty-two out of 72 trait pairs are associated with at least one high-confidence pleiotropic gene (Supplementary Fig. 14b). The highest number of associated pleiotropic genes is observed for ever-asthma and schizophrenia (N=133), followed by ever-asthma & major depressive disorder (N=115), and ever asthma & PTSD (N=106, Fig. 4a). The number of pleiotropic genes associated with ever asthma & ADHD (N=49), ever asthma and bipolar disorder (N=38), and ever-asthma and smoking initiation (N=38) is more modest but still substantial. However, colocalization analysis of pleiotropic genes showed that the fraction of genes harboring signals colocalizing across the two traits was higher (see Methods, Supplementary Fig. 14a) for ever asthma & smoking initiation (96.9%), ever asthma & ADHD (71%), and ever-asthma & major depressive disorder (78%) than for ever asthma & bipolar disorder (41.7%), and for ever-asthma & schizophrenia (12.7%). Thus, ever asthma might share more often the same cis-regulatory mechanisms with smoking initiation, or ADHD, or major depressive disorder than with schizophrenia or bipolar disorder.

**Fig. 4.**
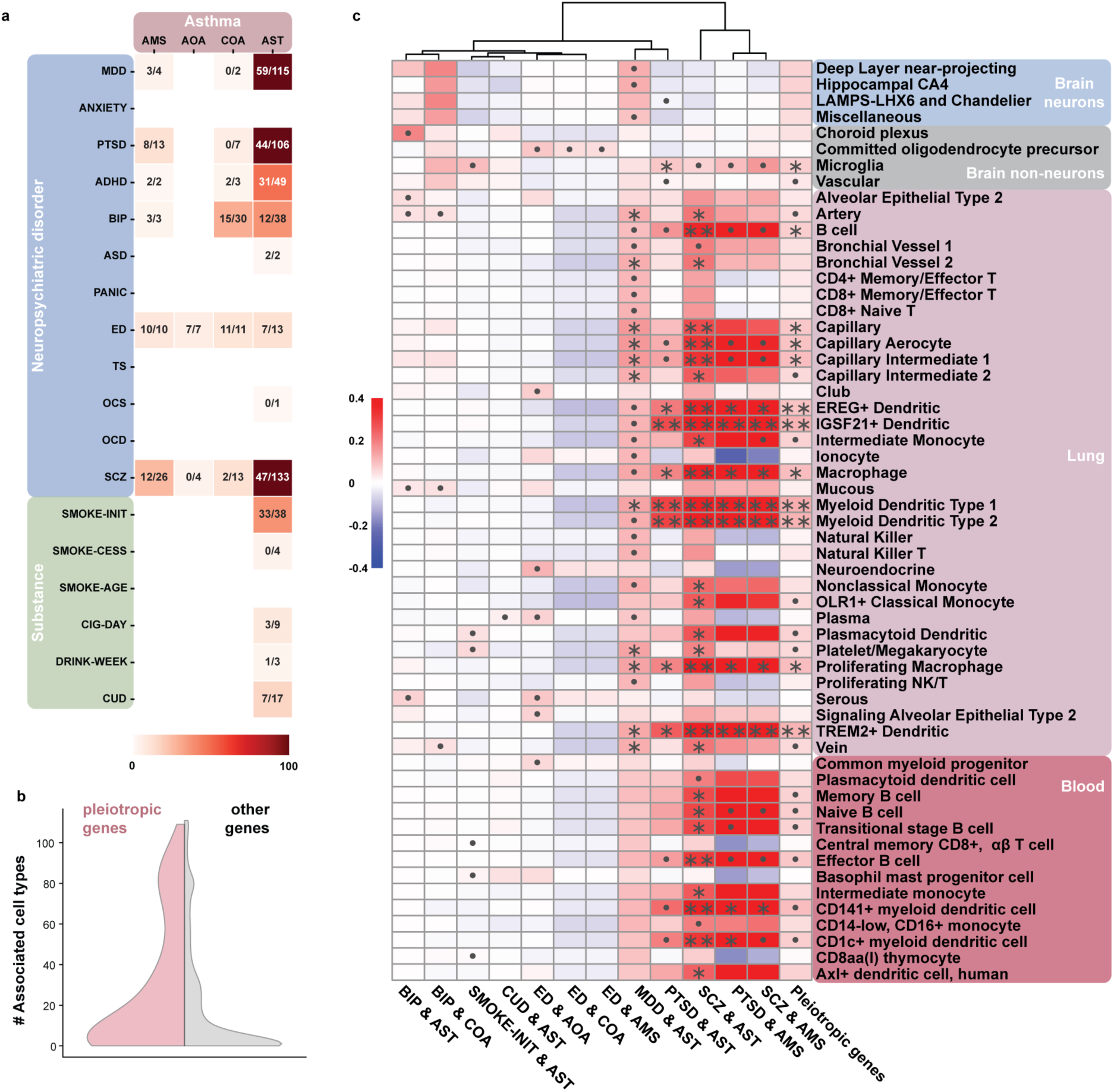
| Pleiotropic genes characterization. **a)** Heatmap showing the number of pleiotropic genes associated with each trait pair. In each cell, the annotation on the left represents the number of high confidence pleiotropic genes, and the annotation on the right represents the total number of pleiotropic genes. The color scale represents the total number of pleiotropic genes. **b)** Violin plot showing the distribution of the number of cell types in which a gene is expressed for high-confidence pleiotropic genes (light pink) and other protein-coding genes (light grey) across brain, blood, and lung. A gene is considered to be expressed in a cell type, if its expression level above the 80^th^ percentile of protein-coding gene expression in the cell type of interest. **c)** Heatmap showing cell-type enrichment scores for the high-confidence gene sets associated with a trait pair across cell types. Positive values (in red) indicate that the expression of the trait-pair gene set is higher than the expression of matching random genes in the cell type. Asterisks indicate significant enrichment (bin-matched permutation test *: *P*_FDR_ < 0.05, and **: *P*_FDR_ < 0.005) and dots indicate nominally significant enrichment (*P* < 0.05). For clarity, cell types with no significant enrichment signals are not shown.

#### Expression patterns of pleiotropic genes in single-cells from brain, lung and blood tissues

We further explored the expression patterns of 178 high-confidence pleiotropic genes across cell-types. We curated and harmonized single-cell RNA-seq datasets from relevant tissues: blood^22^, lung^23^, and brain^24^. We first examined the number of cell-types in which high-confidence pleiotropic genes are expressed compared to other protein coding genes (see Methods). Pleiotropic genes are expressed in more cell types overall and in the blood and lung in particular (Rank-test *P* < 0.008, Fig. 4b, Supplementary Fig. 15a). We then derived a cell-type enrichment score^30^ for combined pleiotropic gene and for each set of high confidence pleiotropic genes associated with a given trait pair (see Methods). Overall, 12 trait pairs were associated with at least one cell type, 57 cell types were associated with at least one trait pair or the combined pleiotropic gene set (8 in the brain, 35 in the lung, and 14 among blood circulating immune cells), yielding 160 associations in total. Enriched cell types varied across trait pairs (Fig. 4c, Supplementary Fig. 16, Supplementary Data 15-16). For instance, ever asthma and schizophrenia or PTSD are primarily associated with myeloid dendritic cells (*P*_FDR_ ≤ 0.00057) and macrophages (*P*_FDR_ ≤ 0.044) of the lung, whereas ever asthma and major depressive disorder are associated with capillary cells, artery and vein of the lung (*P*_FDR_ ≤ 0.041). The combined pleiotropic gene set and ever asthma & PTSD are associated with microglia (*P*_FDR_ ≤ 0.044). Some associations did not survive FDR correction but reached nominal significance, providing complementary interesting biological signals. Ever asthma and major depressive disorder showed nominally significant associations with deep-layer near-projecting, hippocampal CA4, miscellaneous neurons of brain (*P* ≤ 0.039), whereas bipolar disorder and ever asthma showed nominally significant associations with choroid plexus epithelial cells (specialized ciliated cells in the brain that produce cerebrospinal fluid, *P* = 0.028), and mucus-producing cells in the lung (serous, mucous and type 2 alveolar epithelial cells, *P* ≤ 0.027). The diversity of cell-types highlighted by the single cell analysis reflected the diversity of pathways found (immune, neurological, and related to transcription regulation) when performing gene-set enrichment analysis (Supplementary Note 8).

### Immune-related mechanisms, NOTCH signaling, and ciliary dysfunction contribute to the genetic pleiotropy between asthma and neuropsychiatric disorders

We then investigated genes overexpressed in key cell types of interest identified in the previous analysis, focusing on immune cells and microglia, choroid plexus epithelial cells, and neuronal cells. Microglia are resident macrophages of the brain and the main form of active immune defense in the central nervous system, regulating brain development, the maintenance of neuronal networks, and injury repair^31^. We identified genes significantly more expressed in microglia compared to other cell types in the brain (see Methods, Supplementary Data 17-18). Among the 178 identified high confidence pleiotropic genes, 32 were significantly overexpressed in microglia (Supplementary Fig. 17), of which 9 were highly specific to microglia in the brain (i.e. lowly expressed in neurons, Supplementary Fig. 18). Notably, among these 9 genes, 7 are HLA-related genes belonging to the major histocompatibility complex (MHC) class II, which are primarily involved in antigen presentation to CD4⁺ T cells. As thoroughly documented, MHC class II genes are overexpressed in B cells and dendritic cells, including in our dataset (Figs. 5a, b). Beyond the *HLA-D* genes, other immune-related genes are overexpressed in microglia and immune cells across tissues (Figs. 5a, b). For instance, *MICB,* involved in presenting cellular stress signals to cytotoxic cells, was also overexpressed in T cells and NK cells in blood (Fig. 5a). *MICB* can impair the response to rhinovirus in asthma patients when insufficiently secreted^32^ and was associated with psychiatric disorders in a Proteome-Wide Association Study^33^. *ARHGAP15* is expressed to sustained level in microglia in the brain, but also in dendritic cells, macrophages, NK and T cells in the lung and blood (Fig. 5b, Supplementary Fig. 19-20). *ARHGAP15* regulates the polymerization of actin filaments, a key process for macrophage and microglial phagocytosis and migration. This first set of genes highlights mechanisms shared by immune cells in the brain, blood and the lung.

**Fig. 5.**
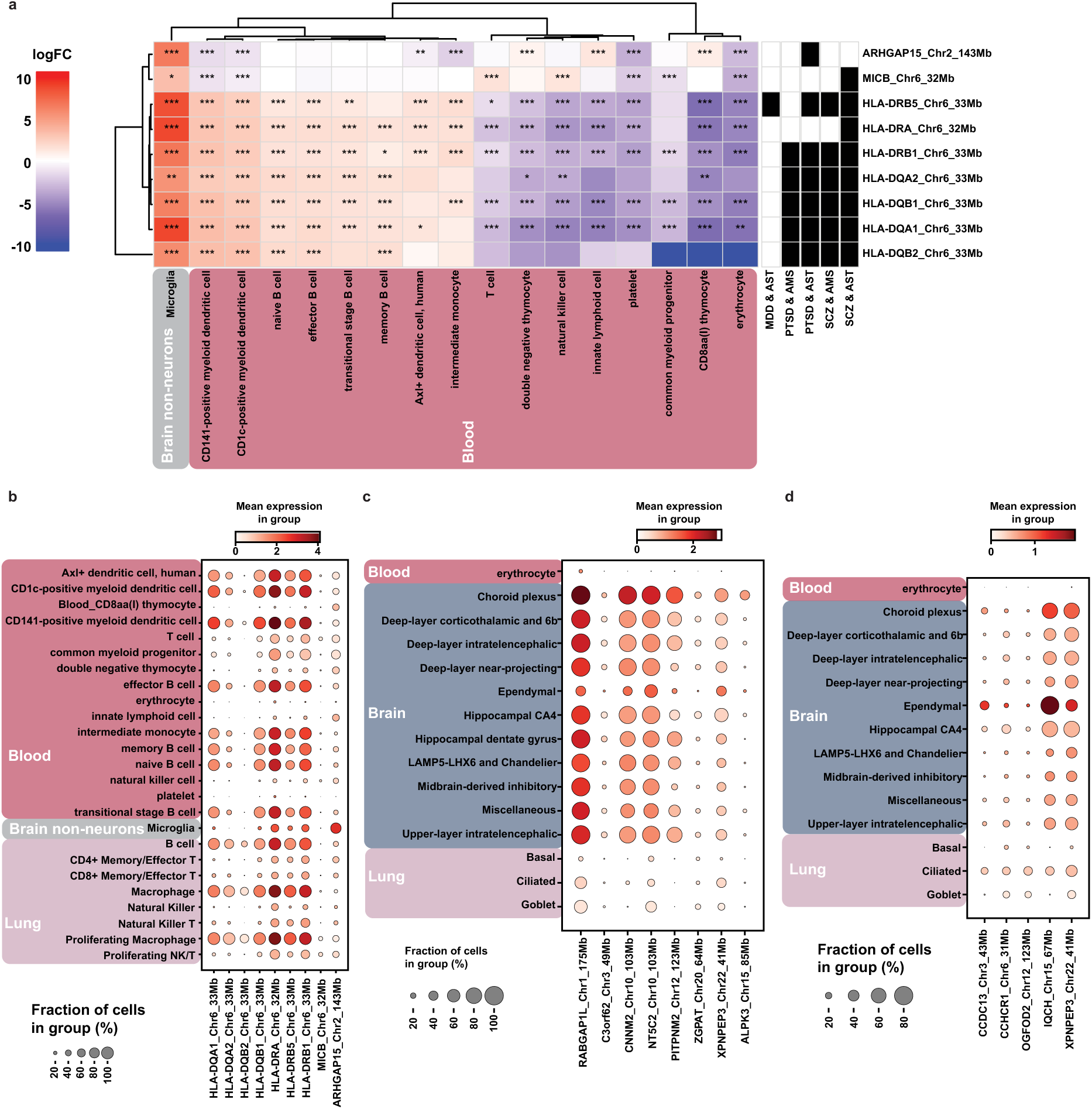
| Differential gene expression analysis and expression of high-confidence genes of interest in microglia, choroid plexus epithelial cells, ependymal cells, and ciliated cells of the lung. **a)** Heatmap showing the log2 fold changes (log2FC) in the expression of immune-related genes of interest for the microglia in each cell type relative to all other cell types within the same tissue. Color intensity represents the log2 fold change of gene expression in each cell type. Statistical significance is indicated by asterisks following Bonferroni correction (nominal *P* × (178 × 107): *** *P* < 0.0005, ** *P* < 0.005, * *P* < 0.05. **b-d)** Dot plots showing the log10 transformed mean expression of genes by cell type among expressing cells as dot color and the fraction of cells expressing each gene as the dot size. **b)** Expression of genes of interest for microglia in immune cells. **c)** Expression of genes of interest for the choroid plexus in ciliated cells, and neurons. **d)** Expression of genes of interest for the ciliated cell types (choroid plexus, ependymal, and ciliated cells of the lung) in ciliated cells, and neurons.

Other genes overexpressed in microglia had a more pervasive expression in the brain (Supplementary Fig. 18), including substantial expression in neurons. Among these, we identified four transcription regulator genes (*EP300*, *ZNF592*, *MAML3,* and *RERE*) implicated in neurodevelopmental pathways, and neurological diseases. Moreover, three of them have been linked to rare genetic diseases (identified using OMIM^34^) with extensive neurological impairments (*ZNF592*^35^*, EP300*^36,37^ and *RERE*^38^, see Supplementary Note 10). Although no rare genetic disease has been attributed to *MAML3,* this gene has previously been associated with bronchial hyperresponsiveness^39^ and migraine^40^. Three of these transcription factors (*RERE*^41^, *EP300*, *MAML3*) are regulators of NOTCH signaling, a major signaling pathway in neurodevelopment, but also in microglia activation during inflammation^42,43^, and in T-cell lineage commitment^44^. Beyond their pervasive expression in the brain, these genes are also expressed in common lymphoid progenitors in blood (Supplementary Fig. 19).

Pleiotropic genes associated with ever-asthma and bipolar disorder are preferentially expressed in choroid plexus epithelial cells, a specialized ciliated cell type of the brain. Among the seven genes overexpressed in the choroid plexus epithelium, three are associated with rare genetic diseases (*CNNM2*^45^, *NT5C2*^46^ and *XPNPEP3*^47^, see Supplementary Note 10), and are expressed in a fraction of neurons, ependymal cells, and also in ciliated cells of the lung (Fig. 5c, Supplementary Figs. 21, 23). The role of *XPNPEP3* in ciliary function, together with the expression of these three genes across ciliary cells of both lung and brain structures, suggests potential role of ciliary dysfunction as a shared impairment between the lung and brain. To explore this hypothesis, we examined genes overexpressed in lung ciliary cells and in at least one ciliary cell type of the brain (ependymal or choroid plexus epithelial cells), and identified five candidates: *XPNPEP3*, *IQCH*, *CCDC13*, *CCHCR1*, *OGFOD2*. In the lung, these five genes were expressed mainly in ciliated cells, but also in basal and goblet cells for *CCHCR1*, *OGFOD2* and *XPNPEP3* (Fig. 5d). In the brain, they exhibit a higher expression in ciliated cells (ependymal and choroid plexus epithelial) but were also expressed in a fraction of neurons. Overall, they show a low expression in blood, except in some progenitor immune cells (Supplementary Fig. 22). Among these genes, three have document key function for ciliary function: *XPNPEP3, CCDC13* regulates microtubule growth^48^, *CCHCR1* is required for ciliogenesis^49^. It is of note that we previously identified genes associated with atopy, known to play a major role in asthma, which contribute to ciliary function^50^.

Finally, we investigated the function of *KCNG2*, the only gene overexpressed in a neuronal subtype associated with major depressive disorder & ever asthma (hippocampal CA4 neurons). This potassium channel is also expressed in plasmacytoid dendritic cells (Supplementary Fig. 24) highlighting the pleiotropic functions of certain ion channel across neurons and immune cells.

## Discussion

We performed an integrative analysis across asthma subtypes, neuropsychiatric disorders, and substance use behaviors, assessing the extent of their shared genetic components through an array of complementary approaches (global and local genetic correlations, bivariate MiXeR, and bivariate GWAS). Through the detection of 138 significant global genetic correlations, complemented by bivariate MiXeR analysis, we identified a robust pattern of genetic association between adult-onset asthma, moderate-to-severe asthma, mood disorders (major depressive disorder, anxiety, and PTSD), ADHD, and smoking behaviors. Bivariate GWAS revealed 354 pleiotropic associations of which 11 are novel. The largest number of pleiotropic loci is found between ever asthma, PTSD, and MDD, further supporting a strong shared genetic etiology between asthma and mood disorders. Through the detection of 178 high-confidence pleiotropic genes, we identified varied pathways underlying these associations: immune pathways (antigen presentation via MHC class II, and complement), neurological pathways (dopamine secretion), and signal transduction (the NOTCH pathways). The 57 enriched cell types are also diverse spanning immune cells (e.g. microglia, dendritic cells, and macrophages), neurons, capillary cells, and ciliated cells. Differential gene expression analysis to identify overexpressed genes in enriched cell-types confirmed the relevance of NOTCH signaling, and immune pathways in understanding the shared genetic risk between asthma and neuropsychiatric disorders. This fine-grained annotation further highlighted ciliary dysfunction and ion channels^51^ as an additional pleiotropic mechanism affecting the brain and the lung.

We detected several highly pleiotropic loci along with high-confidence pleiotropic genes within them (Table 1). Among these genes, several have documented mechanisms that could explain their association with asthma and NPDs. We identified a missense variant in the most pleiotropic locus (Chr3-50Mb, associated with ever asthma, PTSD, ADHD, cannabis use disorder, smoking initiation, major depressive disorder) in the *SEMA3F* gene. Semaphorins were first described for their role in axon guidance during the central nervous development, but they have an increasingly recognized role in the immune system^52^. Specifically*, SEMA3F* can trigger the reorganization of F-actin filaments in mature dendritic cells, facilitating their migration^53^. In addition to the identification of *SEMA3F* as a high-confidence pleiotropic gene, we also identify a lower-confidence association of the *PLXNC1* gene, a transmembrane semaphoring receptor, with ever asthma and PTSD. Within the Chr15-91Mb locus (moderate-to-severe asthma, PTSD, major depressive disorder, bipolar disorder, schizophrenia), the *FURIN* gene encodes a proprotein convertase essential for the activation of numerous substrates including *BDNF* (a neurotrophin implicated in schizophrenia pathophysiology), NOTCH receptors, and the anti-inflammatory cytokine TGF-β1^54^. Within the locus Chr3-71Mb, the *FOXP1* is a transcription factor associated with a neurodevelopmental condition^55^ characterized by intellectual disability, but is also implicated in B-cell and T-cell differentiations^56^. These genes highlight additional biological mechanisms of horizontal pleiotropy affecting both the immune and nervous systems.

Despite strong genetic and epidemiological evidence for the association of smoking with asthma and neuropsychiatric disorders, to our knowledge, smoking has never been included in a study on the shared genetic etiology between asthma and NPDs. Although epidemiological reports can vary by country, smoking is overall as prevalent in asthma patients as in the general population^57–59^ despite its causal link to the worsening of symptoms and resistance to inhaled corticosteroid treatment^58^. Smoking is also associated with later asthma onset with a stronger effect in women^57,58^. Smoking rates are higher in patients with mental illness, with particularly elevated odds ratios in schizophrenia, bipolar disorder, and PTSD^60,61^. Smoking behaviors and psychiatric disorders were also correlated genetically in previous reports^16^ as confirmed by our study. We assessed the potential mediating effect of smoking on the genetic association between asthma and NPDs by including smoking behaviors in our analysis. Although we found a substantial overlap between ever asthma, adult-onset asthma, moderate-to-severe asthma and smoking, smoking cannot fully account for the shared genetic architecture between asthma and NPDs. Indeed, the estimated number of causal variants and the number of pleiotropic loci shared between asthma and NPDs is larger than the number shared between asthma and smoking behaviors.

Consistent with a previous study^11^, the genetic correlations observed between asthma subtypes, bipolar disorder and schizophrenia are moderate. At the locus level, we observed a larger genetic overlap: more than 10 loci are associated with an increased risk for ever asthma, and schizophrenia or bipolar depression. Bivariate MiXeR analysis revealed a large overlap between ever asthma and bipolar disorder and ever asthma and schizophrenia, but with a fraction of concordant genetic effect sign of approximately 50%. At the epidemiological level, while schizophrenia and bipolar disorder are robustly associated with asthma, several factors modulate these associations. Asthma is more frequent in patients with bipolar disorder comorbid with migraines, a subtype of bipolar disorder more prevalent in women^62^. Moreover, female patients with asthma and schizophrenia or bipolar disorder have an earlier onset of positive symptoms (i.e. manic episode, or psychosis), but this effect is not observed in male patients^63^. Together these observations suggest a more complex relationship between asthma, bipolar disorder and schizophrenia that might be modulated by clinical subtypes or might be sex-dependent. A deeper investigation of the shared genetic architecture between schizophrenia, bipolar disorder, and asthma, including finer clinical subtypes, asthma immune-related traits (e.g., IgE levels, and eosinophil count), and GWAS stratified by sex could help to clarify this picture.

Genetic correlations between childhood-onset asthma and neuropsychiatric traits are moderate, underscoring the importance of including asthma subtypes when studying the relationship between asthma and NPDs. In contrast to Zhu et al^12^, we found several significant global genetic correlations between childhood-onset asthma and neuropsychiatric disorders, likely attributable to the larger sample sizes of GWAS included in this study. Childhood-onset asthma is associated with major depression disorder, and PTSD confirming a previous report of the genetic liability of major depression disorder being modestly associated with childhood-onset asthma^25^. We also observed a nominally significant global genetic correlation between childhood-onset asthma and bipolar disorder (*P*=0.03), and detected several pleiotropic loci associated with both traits. Epidemiological reports on the associations between early-onset asthma and subsequent bipolar disorder are contrasted: two studies^8,64^ reported a significant increased risk for both disorders whereas a study focusing on men does not find a convincing link between these two traits^65^. Therefore, the association between childhood-onset asthma and bipolar disorder may be clinical subtype- or sex-dependent, a question that may be addressed in future studies.

Our study has limitations. The limited effective sample sizes of the studies on childhood-onset asthma, adult-onset asthma, and moderate-to-severe asthma GWAS restricted the detections of pleiotropic loci. Thus, most of the pleiotropic loci are associated with ever asthma preventing from distinguishing between pathways specifically associated with an asthma subtype in the functional analysis. Computing polygenic risk scores stratified by biological pathways^66^ would allow the assessment of the association of pathways identified in this study with each asthma subtypes, clarifying their contribution to asthma pathophysiology. Our analysis focused on GWAS of European ancestry, and further studies of non-European ancestries will be necessary. At the date of the preparation of this manuscript, existing data in non-European ancestries is only available in a limited number of traits covered by this analysis. We did not perform fine-mapping despite the existence of multi-trait fine-mapping methods^18,67,68^. Fine-mapping methods may perform poorly when applied to GWAS meta-analysis summary statistics, as heterogeneity in sample size, phenotyping, genotyping, and imputation across contributing cohorts can distort the concordance between meta-analysis association statistics and the reference LD structure, leading to miscalibrated fine-mapping results^69^. Finally, the tissues used in single cell data were not obtained from patients with a neuropsychiatric disorder or affected by asthma and might not reflect cell type states observed in disease (e.g. microglia subtypes associated to schizophrenia^70^).

In conclusion, our integrative study found a large genetic overlap between mood disorders and asthma, which is stronger for adult-onset asthma and moderate-to-severe asthma. The shared genetic etiology between bipolar or schizophrenia disorder and asthma is more moderate and may be clinical subtype dependent. We found no evidence of a shared genetic etiology between asthma and obsessive-compulsive disorders. Although smoking is involved in the shared genetic etiology between asthma and NPDs, it accounts for only a minority of the pleiotropic loci. Through a comprehensive functional analysis, we pinpointed several biological mechanisms likely to exert horizontal pleiotropy across the immune and nervous system such as antigen presentation, NOTCH signaling, ciliary dysfunction, and ion channels. These findings might help identify new treatment strategies for patients with asthma co-occuring with neuropsychiatric disorders.

## Methods

### GWAS summary statistics included in the present study

We curated 24 GWAS summary statistic datasets (Supplementary Data 1) corresponding to the most recent studies available on traits of interest at the time of the manuscript preparation (spring 2025). We included four distinct GWAS summary statistic datasets of asthma subtypes: ever asthma from the Global Biobank Meta-analysis Initiative (GBMI), which aggregates data from 14 biobanks^71^; childhood-onset asthma (age of onset ⩽ 12 years) and adult-onset asthma (age of onset ⩾ 26 years)^12^ both based on the UK Biobank data; and moderate-to-severe asthma^72^. For sensitivity analyses, we included two additional GWAS on childhood-onset asthma (age of onset ⩽ 19 years) and on adult-onset asthma (age of onset ⩾ 20 years)^73^ also based on UK Biobank data, but with different age of asthma onset thresholds.

For neuropsychiatric disorders, we included GWAS summary statistics for 12 conditions based on Psychiatric Genomics Consortium (PGC) data: major depressive disorder (MDD)^74^, panic disorder (PANIC)^75^, anxiety disorder (ANXIETY)^76^, attention-deficit/hyperactivity disorder (ADHD)^77^, autism spectrum disorder (ASD)^78^, bipolar disorder (BIP)^79^, Post-traumatic stress disorder (PTSD)^80^, Tourette Syndrome (TS)^81^, obsessive-compulsive symptoms (OCS)^82^, Obsessive-compulsive disorder (OCD)^83^, eating disorders (ED)^84^ and schizophrenia (SCZ)^85^. Note that for schizophrenia and eating disorder, the most recent studies did not pass all the QC steps of the LD Score regression pipeline (Supplementary Note 12). Hence, we worked with the penultimate studies available on these traits. We included six GWAS summary statistics on substance use behaviors. Five smoking and drinking related traits were retrieved from LIU et al^16^: smoking initiation (i.e., having ever regularly smoked), smoking cessation (i.e., being a former smoker), age at smoking initiation, cigarettes smoked per day, and drinks per week. These were complemented by cannabis use disorder^86^ from PGC (Psychiatric Genomics Consortium).

### Quality control, harmonization, and imputation of GWAS summary statistics

All GWAS summary statistics were harmonized using the Joint Analysis of Summary Statistics (JASS) pre-processing pipeline^87^. This process involved: (1) mapping heterogeneous GWAS column names to standardized ones; (2) selecting variants present in the reference panel (in this case, hg38 European panel for common variants, MAF>1%, based on 1000 Genomes Project Phase 3 data); (3) aligning the effect alleles from the GWAS datasets with the reference panel; (4) inferring sample sizes for each variant based on MAF, standard deviation, and genetic effect; (5) filtering variants with a sample size lower than 70% of the 90^th^ percentile of sample size across variants; and (6) computing a standardize genetic effect size (Z-score).

As all GWAS did not report the same sets of variants, we performed imputation of missing summary statistics as implemented in RAISS^88^, allowing us to increase the number of variant-trait pairs with a Z-score that will be available for multi-trait pairwise GWAS. The approach leverages linkage disequilibrium between variants to predict missing *Z*-score from the available Z-scores of neighboring variants. The *raiss* command was run with the following parameter setting: --R2-threshold=0.6, --eigen-threshold=0.05, --minimum-ld=5. The imputation quality was tested for all traits with the subcommand *raiss performance-grid-search.* The highest median imputation error across trait (median(|Z^observed^ – Z^imputed^|) = 0.16) was observed for anxiety disorder, indicating an overall satisfactory imputation quality (Supplementary Data19).

### Global and local genetic correlations, and genetic overlap

We estimated the global genetic correlations across all trait pairs using LD Score Regression (LDSC) Python package^19^. Input data consisted of our harmonized GWAS files and of LD scores computed for our reference panel (European hg38, based on 1000 Genomes Project Phase 3 data; https://zenodo.org/record/8096589).

We estimated the local genetic correlations across all trait pairs in 2,108 quasi LD-independent genomic regions^89^ using LAVA^20^. For each trait pair, we selected regions in which both traits showed significant local variant-based heritability (*P* < 0.05) and computed their local genetic correlations. Sample overlap between trait pairs (input required by LAVA) was estimated by LD-score regression intercepts. P-values for both global and local genetic correlations were adjusted using the Benjamini-Hochberg false discovery rate procedure (*P*_FDR_ < 0.05).

To estimate the genetic overlap among asthma subtypes, neuropsychiatric traits and substance use behaviors irrespective of effect direction, we used the univariate and bivariate causal mixture models for GWAS summary statistics implemented in MiXeR. Bivariate MiXeR^21^ is a Bayesian model that decomposes the genetic architecture of a trait pair into trait-specific and shared polygenic components and estimates the global genetic correlation based on these polygenic structures. Bivariate MiXeR provides estimates for the polygenicity of shared and specific polygenic components (reported using the number of causal variants in thousand), the genetic correlation between the two traits, and the fraction of shared causal variants with concordant effect signs. Following authors’ recommendation, the effective sample size for binary traits was calculated as Neff = N × 4 × p × (1 - p), where N denotes the total sample size and p represents the proportion of cases in the study sample.

### Multi-trait pairwise GWAS analysis

To detect pleiotropic variants, we performed multi-trait pairwise GWAS using the omnibus test for each trait pair between asthma subtypes (ever asthma, childhood-onset asthma, adult-onset asthma and moderate-to-severe asthma) and all other traits (neuropsychiatric disorders, and substance use behaviors). For each trait pair, we selected variants meeting the following criteria: 1) omnibus *P* < 5x10^-8^/72 (Bonferroni correction, 72 trait pairs); 2) univariate *P*-values for both traits below 10^-5^; and 3) concordant genetic-effect sign for both traits (except for pairs containing age-at-smoking initiation, for which we selected opposed effect signs as a smaller age at smoking initiation corresponds to a more severe phenotype). As an additional control step, we excluded omnibus significant variants that were not surrounded by variants with significant or near significant omnibus *P*-values (*P* < 10^-6^) in the same genomic region. Then, for each trait pair, we identified lead variants using the PLINK clumping procedure applied to the omnibus *P*-values (parameters: --clump-p1 5x10^-8^/72, --clump-p2 10^-5^/72, --clump-r2 0.8, --clump-kb 250). We refer to these variants as lead pleiotropic variants. We considered these associations to be novel if the lead pleiotropic variant was not in LD (r²>0.1, within 250kb) with any single-trait GWAS significant variants of the corresponding trait pair.

### Colocalization analysis

We conducted a colocalization test, as implemented in coloc^90^, to assess if pleiotropic variants are likely to reflect the same causal variant for a given trait pair. For each lead pleiotropic variant of a given trait pair, we extracted univariate association results (Z-scores) of both traits for variants within a ±250 kb window to perform the colocalization test. Coloc estimates the posterior probabilities of five hypotheses: **H0**: No association with either trait; **H1**: association with trait 1 only; **H2**: association with trait 2 only; **H3**: association with both traits driven by two independent variants; **H4**: association with both traits driven by one causal variant. We classified variants with pp.H4 ≥ 0.8 as colocalized pleiotropic variants, while others were classified as non-colocalized pleiotropic variants.

### Annotation of pleiotropic lead variants for consequences, biotypes, and functional categories

We annotated pleiotropic variants using the Ensembl Variant Effect Predictor (VEP), which predicts the effects of variants on gene transcripts, protein sequences, and regulatory regions (https://www.ensembl.org/info/docs/tools/vep). We considered only the canonical transcript to avoid redundant annotation of the same variants. In addition, we retrieved the functional ranks as defined in RegulomeDB^28^, which provide a proxy of how likely a variant is to be functional by looking overlaps with transcription factor binding sites, Dnase peaks, and eQTLs.

### Assignation of genetic variants to genes

To assign pleiotropic variants to candidate effector genes, we performed positional mapping and eQTLs mapping in relevant tissues. Firstly, we assigned the lead pleiotropic variants to the closest gene within ±50 kb window size by using the Ensembl Human genes (GRCh38.p14, release114, https://www.ensembl.org/) and focused on the 19,156 protein coding genes on Chromosome 1-22. Secondly, we searched for effector genes through cis-eQTLs using the Genotype Tissue Expression (GTEx) database (GTEx_Analysis_v10_eQTL, https://gtexportal.org/home/downloads) containing significant variant–gene expression associations (*P*_FDR_ < 0.05) across 50 human tissues. We focused on tissues that were relevant to our traits of interest, including 13 brain-related tissues, lung tissue and whole-blood. For each lead pleiotropic variant, we gathered the results of these 13 brain tissues together and chose the one with the most significant *P*-values to represent brain tissue. To lookup for the overlap between eQTLs and our lead pleiotropic variants, we additionally considered all LD proxy variants (distance≤250kb and LD r2≥0.8 with the lead pleiotropic variants) which are in the cis-regulatory element as defined by the ENCODE Candidate Cis-Regulatory Elements (cCREs) database (https://genome.ucsc.edu/cgi-bin/hgTables). This database provides a total of 926,535 elements of 5 types: promoter-like signature, proximal enhancer-like signature, distal enhancer-like signature, DNase-H3K4me3, CTCF-only. For both positional mapping and eQTLs mapping genes, if the original lead pleiotropic variant passed the colocalization test (pp.H4≥0.8), we noted them as colocalized-pleiotropic genes, while the others were kept as candidate pleiotropic genes.

### Gene-based analysis

We identified additional candidate pleiotropic genes using fastBAT^29^ which computes aggregated *P* value of variants located within ±50 kb of a gene, using GWAS summary statistics for a single trait and the linkage disequilibrium (LD) structure among these variants. For each trait pair, we identified the genes whose aggregated *P-*values are significant (*P* < 2.5x10^-6^) for both traits. To ensure that each pleiotropic gene exhibited consistent effect directions across both traits, we collected variants located within ±50 kb of each gene and calculated: (1) the proportion of all common variants showing the same effect direction, and (2) the proportion of variants in both traits (GWAS *P* < 10^-5^ for both traits) showing the same effect direction. Genes with both proportions greater than 0.5 were retained for further analysis. Then, for these genes, we extracted univariate association results (Z-scores) for all variants within ±50 kb of the gene region for both traits and performed a colocalization test to determine whether the association signals were driven by the same variants in both traits. We considered genes with pp.H4 ≥ 0.8 as colocalized (Supplementary Data 20).

### High-confidence pleiotropic genes

Finally, we identified pleiotropic genes for each trait pair using five methods: positional mapping, cis-eQTL in brain, cis-eQTL in lung, cis-eQTL in whole_blood, and burden test as implement in fastBAT^29^. By integrating results from these five methods, we identified 315 candidate pleiotropic genes across 27 trait pairs. To improve their reliability, we retained, for each trait pair, the pleiotropic genes that either (i) passed the colocalization test (pp.H4 ≥ 0.8) in at least one method, or (ii) were identified by at least three methods regardless of colocalization. After filtering, we obtained 178 high-confidence pleiotropic genes across 22 trait pairs, and all subsequent analyses were based on these refined pleiotropic genes.

### Gene set enrichment analysis

For each high-confidence pleiotropic gene set associated with a given trait pair, we performed pathway enrichment analyses using databases from MSigDB v2025.1 (GSEA, https://www.gsea-msigdb.org/), including GO Biological Process (GOBP), GO Cellular Component (GOCC), WikiPathways, Reactome, Human Phenotype Ontology (HPO), and KEGG pathways from BioServices. This enrichment was assessed using a hypergeometric test, with protein coding gene as background gene set. P-values were adjusted using the Benjamini– Hochberg FDR false discovery rate procedure, for each trait pair and each pathway database.

### Single cell RNA-seq: expression breadth

To identify cell-types involved in asthma and neuropsychiatric traits comorbidities, we curated and harmonized single-cell RNA-seq datasets of three relevant tissues: peripheral blood including circulating immune cells^22^, adult lung^23^ and adult brain including neurons and non neurons^24^ (Supplementary Data 21).

To assess whether high-confidence pleiotropic genes are expressed across a broader range of cell types than randomly selected protein-coding genes, we applied the following approach. For each protein-coding gene and cell type, we aggregated raw counts across all cells belonging to that cell type. Within each cell type, genes ranking in the top 20% by aggregated raw counts were classified as highly expressed. For each tissue separately and across all three tissues combined, we defined the expression breadth of each gene as the number of cell types in which it was classified as highly expressed. We then performed one-sided Wilcoxon rank-sum tests to determine whether high-confidence pleiotropic genes exhibit significantly greater expression breadth than other protein-coding genes. Additionally, we calculated the proportions of high-confidence pleiotropic genes and other protein-coding genes classified as highly expressed in at least one cell type within each of the three tissues. P-values and odds ratio were assessed using Fisher’s exact test.

### Single cell RNA-seq: cell-type enrichment

To determine cell types preferentially associated with each trait pair, we computed cell-type enrichment for each gene set associated with a trait pair and a combined gene set including high confidence pleiotropic genes. We first preprocessed raw counts using the python package Scanpy^30^, including cell-wise normalization, log-transformation, and aggregation of the mean expression levels by gene and cell type. We then calculated cell-type enrichment scores for pleiotropic gene sets associated with each trait pair (n_bins=20) using scanpy.tl.score_genes function. To ensure the reliability of the results, we considered only gene sets comprising of at least five genes. We then computed the significance of cell-type enrichment for each gene set using a bin-matched permutation test. Following the same binning approach in scanpy.tl.score_genes, for each tissue, we grouped the protein-coding genes into 20 bins based on their mean expression levels across all cell types. During the permutation procedure, control genes were randomly sampled from the same expression bin as the pleiotropic genes. Each permutation test was repeated 100,000 times to obtain robust *P-*values, which were subsequently adjusted using the Benjamini–Hochberg false discovery rate procedure.

### Single cell RNA-seq: cell-type-specific differential expression analysis

To detect genes overexpressed in a given cell type, we performed differential gene expression analysis across all cell types within each tissue using glmGamPoi^91^. We constructed a pseudo-bulk expression matrix (genes × samples), where each sample represented a unique donor–cell type combination, obtained by aggregating raw counts across all cells of the same cell type from the same donor. We retained all protein-coding genes with at least 10 raw counts in more than 5% of samples. After filtering, the peripheral blood dataset contained 11,127 genes and 4,012 samples, the adult lung dataset contained 12,520 genes and 114 samples, and the brain dataset contained 17,590 genes and 122 samples.

For each cell type, samples belonging to that cell type (across all donors) were designated as cases and all other samples as controls. Differential expression analysis was then performed using glmGamPoi R package with the normed_sum normalization method. To minimize the impact of inter-donor variability, the donor identity was included as a covariate in the model. For each gene, we estimated the expression fold change (log2FC) and the corresponding *P-*value. A positive log2FC indicates that the gene has higher expression in the target cell type relative to all other cell types within the same tissue. We applied a Bonferroni-corrected significance threshold of *P* < 0.05/(178*107), where 178 is the number of high confidence pleiotropic genes, and 107 is the number of tested cell types.

## Data availability

All analyses in this study were based on publicly available data. The sources of GWAS summary statistics are listed in Supplementary Data 1. The GTEx eQTL v10 was downloaded from the GTEx Portal (https://www.gtexportal.org/home/downloads/adult-gtex/qtl#qtl-gtex_analysis_v10-single-tissue_cis-qtl_data). Cis-regulatory element annotation was downloaded from ENCODE3 cCREs https://genome.ucsc.edu/cgi-bin/hgTables. The gene annotation ENSEMBL v114 was download from https://www.ensembl.org/. The GWAS Catalog all associations v1.0.2 was retrieved from the https://www.ebi.ac.uk/gwas/docs/file-downloads/. The 1000 genomes reference of European ancestry was downloaded from https://zenodo.org/records/11193775. The RegulomeDB variant annotation was downloaded from https://regulomedb.org/regulome-search/. The pathway dataset was retrieved from MSigDB v2025.1 (https://www.gsea-msigdb.org/gsea/msigdb) and BioServices (KEGG pathway, https://bioservices.readthedocs.io/en/latest/kegg_tutorial.html). The brain single cell analysis dataset was downloaded from https://github.com/linnarsson-lab/adult-human-brain. The lung single cell analysis dataset was downloaded from https://www.synapse.org/Synapse:syn21625095. The blood single cell analysis was downloaded from https://cellxgene.cziscience.com/collections/77f9d7e9-5675-49c3-abed-ce02f39eef1b.

## Code availability

JASS pipeline for GWAS summary statistics harmonization, preprocessing, imputation, computation of genetic correlations and multi-trait pairwise analysis was from https://gitlab.pasteur.fr/statistical-genetics/jass_suite_pipeline. Other software in this study included PLINK v1.9 (https://www.cog-genomics.org/plink/1.9/), LAVA v0.1.0 (https://github.com/josefin-werme/LAVA), MiXeR v1.3.0 (https://github.com/precimed/mixer), Coloc v5.2.3 (https://chr1swallace.github.io/coloc/), fastBAT of GCTA v1.94.1 (https://yanglab.westlake.edu.cn/software/gcta/#fastBAT), Scanpy v1.11.5 (https://scanpy.scverse.org/), glmGamPoi v1.10.2 (https://github.com/const-ae/glmGamPoi). Custom code can be found in https://github.com/Haibo-Huang98/asthma_NPDS_SUBs.

## Acknowledgments

We would like to thank the members of the Genomic Epidemiology of Multifactorial Diseases Group at the HealthFex INSERM Unit and the Statistical Genetics Unit at the Institut Pasteur for their constructive feedback on our preliminary presentations of this work. We would also like to thank Michael Cho for his insightful comments and feedback on an earlier presentation of this work.

## References

1. Vos, T. et al. Global burden of 369 diseases and injuries in 204 countries and territories, 1990–2019: a systematic analysis for the Global Burden of Disease Study 2019. The Lancet 396, 1204–1222 (2020).

2. Koppelman, G. H. et al. Genetic and environmental risk factors for asthma: towards prevention. Lancet Respir. Med. 13, 1011–1025 (2025).

3. Zhong, X. et al. Integration of functional genomics and statistical fine-mapping systematically characterizes adult-onset and childhood-onset asthma genetic associations. Genome Med. 17, 35 (2025).

4. Xu, X. et al. Association between allergic diseases and mental health conditions: An umbrella review. J. Allergy Clin. Immunol. 155, 701–713 (2025).

5. Allgire, E., McAlees, J. W., Lewkowich, I. P. & Sah, R. Asthma and posttraumatic stress disorder (PTSD): Emerging links, potential models and mechanisms. Brain. Behav. Immun. 97, 275–285 (2021).

6. Cortese, S. et al. Association between attention deficit hyperactivity disorder and asthma: a systematic review and meta-analysis and a Swedish population-based study. Lancet Psychiatry 5, 717–726 (2018).

7. Lehto, K., Pedersen, N. L., Almqvist, C., Lu, Y. & Brew, B. K. Asthma and affective traits in adults: a genetically informative study. Eur. Respir. J. 53, 1802142 (2019).

8. Liu, X. et al. Bidirectional Associations Between Asthma and Types of Mental Disorders. J. Allergy Clin. Immunol. Pract. 11, 799–808.e14 (2023).

9. Li, C. et al. Functional Two-Way Crosstalk Between Brain and Lung: The Brain–Lung Axis. Cell. Mol. Neurobiol. 43, 991–1003 (2023).

10. Dill-McFarland, K. A. et al. Molecular pathways underlying lung-brain axis signaling in asthma: Relevance for psychopathology and neuroinflammation. J. Allergy Clin. Immunol. 153, 111–121 (2024).

11. Lu, Z.-A. et al. Genetic overlap of severe psychiatric disorders with lung function and asthma suggests shared biological mechanisms. Brain. Behav. Immun. 129, 305–317 (2025).

12. Zhu, Z. et al. Shared genetics of asthma and mental health disorders: a large-scale genome-wide cross-trait analysis. Eur. Respir. J. 54, 1901507 (2019).

13. Mathew, A. R., Hogarth, L., Leventhal, A. M., Cook, J. W. & Hitsman, B. Cigarette smoking and depression comorbidity: systematic review and proposed theoretical model: Smoking and depression. Addiction 112, 401–412 (2017).

14. de Leon, J. & Diaz, F. J. A meta-analysis of worldwide studies demonstrates an association between schizophrenia and tobacco smoking behaviors. Schizophr. Res. 76, 135–157 (2005).

15. Van Amsterdam, J., Van Der Velde, B., Schulte, M. & Van Den Brink, W. Causal Factors of Increased Smoking in ADHD: A Systematic Review. Subst. Use Misuse 53, 432–445 (2018).

16. Liu, M. et al. Association studies of up to 1.2 million individuals yield new insights into the genetic etiology of tobacco and alcohol use. Nat. Genet. 51, 237–244 (2019).

17. Grotzinger, A. D. et al. Mapping the genetic landscape across 14 psychiatric disorders. Nature 649, 406–415 (2026).

18. Jee, Y. H. et al. Dissecting pleiotropy to gain mechanistic insights into human disease. Nat. Rev. Genet. 27, 292–305 (2026).

19. Bulik-Sullivan, B. et al. An atlas of genetic correlations across human diseases and traits. Nat. Genet. 47, 1236–1241 (2015).

20. Werme, J., Van Der Sluis, S., Posthuma, D. & De Leeuw, C. A. An integrated framework for local genetic correlation analysis. Nat. Genet. 54, 274–282 (2022).

21. Frei, O. et al. Bivariate causal mixture model quantifies polygenic overlap between complex traits beyond genetic correlation. Nat. Commun. 10, 2417 (2019).

22. Gong, Q. et al. Multi-omic profiling reveals age-related immune dynamics in healthy adults. Nature 648, 696–706 (2025).

23. Travaglini, K. J. et al. A molecular cell atlas of the human lung from single-cell RNA sequencing. Nature 587, 619–625 (2020).

24. Siletti, K. et al. Transcriptomic diversity of cell types across the adult human brain. Science 382, eadd7046 (2023).

25. Liu, X. et al. Genetic liability to major depression and risk of childhood asthma. Brain. Behav. Immun. 89, 433–439 (2020).

26. Han, Y. et al. Genome-wide analysis highlights contribution of immune system pathways to the genetic architecture of asthma. Nat. Commun. 11, 1776 (2020).

27. Vuckovic, D. et al. The Polygenic and Monogenic Basis of Blood Traits and Diseases. Cell 182, 1214–1231.e11 (2020).

28. Boyle, A. P. et al. Annotation of functional variation in personal genomes using RegulomeDB. Genome Res. 22, 1790–1797 (2012).

29. Bakshi, A. et al. Fast set-based association analysis using summary data from GWAS identifies novel gene loci for human complex traits. Sci. Rep. 6, (2016).

30. Wolf, F. A., Angerer, P. & Theis, F. J. SCANPY: large-scale single-cell gene expression data analysis. Genome Biol. 19, 15 (2018).

31. Colonna, M. & Butovsky, O. Microglia Function in the Central Nervous System During Health and Neurodegeneration. Annu. Rev. Immunol. 35, 441–468 (2017).

32. Telcian, A. G. et al. Soluble Major Histocompatibility Complex Class I-Related Chain B Molecules Are Increased and Correlate With Clinical Outcomes During Rhinovirus Infection in Healthy Subjects. Chest 146, 32–40 (2014).

33. Gong, W. et al. Genomics-driven integrative analysis highlights immune-related plasma proteins for psychiatric disorders. J. Affect. Disord. 370, 124–133 (2025).

34. Amberger, J. S., Bocchini, C. A., Scott, A. F. & Hamosh, A. OMIM.org: leveraging knowledge across phenotype–gene relationships. Nucleic Acids Res. 47, D1038–D1043 (2019).

35. Nicolas, E. et al. CAMOS, a nonprogressive, autosomal recessive, congenital cerebellar ataxia, is caused by a mutant zinc-finger protein, ZNF592. Eur. J. Hum. Genet. 18, 1107–1113 (2010).

36. Maripuri, D. P., Gold, J., Gold, N. & Strong, A. A New EP300-Related Syndrome With Prominent Developmental and Immune Phenotypes. Am. J. Med. Genet. A. 197, e64050 (2025).

37. Menke, L. A. et al. Further delineation of an entity caused by CREBBP and EP300 mutations but not resembling Rubinstein–Taybi syndrome. Am. J. Med. Genet. A. 176, 862–876 (2018).

38. Jordan, V. K. et al. Genotype–phenotype correlations in individuals with pathogenic RERE variants. Hum. Mutat. 39, 666–675 (2018).

39. Nieuwenhuis, M. a. E., et al. PTTG1IP and MAML3, novel genomewide association study genes for severity of hyperresponsiveness in adult asthma. Allergy 72, 792–801 (2017).

40. Gasperi, M. et al. A multi-ancestry meta genome-wide association study of migraine among veterans: associations with traumatic brain injury, depression, and post-traumatic stress disorder. Mol. Psychiatry 31, 2660–2674 (2026).

41. Vilhais-Neto, G. C. et al. Rere controls retinoic acid signalling and somite bilateral symmetry. Nature 463, 953–957 (2010).

42. Wei, Z. et al. Notch Activation Enhances the Microglia-Mediated Inflammatory Response Associated With Focal Cerebral Ischemia. Stroke 42, 2589–2594 (2011).

43. Grandbarbe, L. et al. Notch signaling modulates the activation of microglial cells. Glia 55, 1519–1530 (2007).

44. Radtke, F., Fasnacht, N. & MacDonald, H. R. Notch Signaling in the Immune System. Immunity 32, 14–27 (2010).

45. Arjona, F. J. et al. CNNM2 Mutations Cause Impaired Brain Development and Seizures in Patients with Hypomagnesemia. PLOS Genet. 10, e1004267 (2014).

46. Novarino, G. et al. Exome Sequencing Links Corticospinal Motor Neuron Disease to Common Neurodegenerative Disorders. Science 343, 506–511 (2014).

47. O’Toole, J. F. et al. Individuals with mutations in *XPNPEP3*, which encodes a mitochondrial protein, develop a nephronophthisis-like nephropathy. J. Clin. Invest. 120, 791–802 (2010).

48. Colin, E. et al. Loss-of-Function Mutations in *WDR73* Are Responsible for Microcephaly and Steroid-Resistant Nephrotic Syndrome: Galloway-Mowat Syndrome. Am. J. Hum. Genet. 95, 637–648 (2014).

49. Zhang, J., Wu, Q., Zhang, S., Sze, S. C. W. & Li, C. CCHCR1 links P-body proteins to the centrosome and is required for ciliogenesis through interacting with OFD1 and PCM1. Cell. Mol. Biol. Lett. 30, 103 (2025).

50. Sugier, P.-E. et al. A novel role for ciliary function in atopy: ADGRV1 and DNAH5 interactions. J. Allergy Clin. Immunol. 141, 1659–1667.e11 (2018).

51. Dardani, C. et al. Revisiting the Links Between Asthma and the Psychosis Spectrum: shared molecular mechanisms. Biol. Psychiatry 10.1016/j.biopsych.2026.07.023 (2026) doi:10.1016/j.biopsych.2026.07.023.

52. Thomas, R. & Yang, X. Semaphorins in immune cell function, inflammatory and infectious diseases. *Curr*. Res. Immunol. 4, 100060 (2023).

53. Curreli, S., Wong, B. S., Latinovic, O., Konstantopoulos, K. & Stamatos, N. M. Class 3 semaphorins induce F-actin reorganization in human dendritic cells: Role in cell migration. J. Leukoc. Biol. 100, 1323–1334 (2016).

54. Zhang, Y. et al. The emerging role of furin in neurodegenerative and neuropsychiatric diseases. Transl. Neurodegener. 11, 39 (2022).

55. Lozano, R. et al. FOXP1 syndrome: a review of the literature and practice parameters for medical assessment and monitoring. J. Neurodev. Disord. 13, 18 (2021).

56. Zaiss, D. M. W. & Coffer, P. J. Forkhead box transcription factors as context-dependent regulators of lymphocyte homeostasis. Nat. Rev. Immunol. 18, 703–715 (2018).

57. Kamga, A. et al. Asthma and smoking: A review. Respir. Med. Res. 82, 100916 (2022).

58. Polosa, R. & Thomson, N. C. Smoking and asthma: dangerous liaisons. Eur. Respir. J. 41, 716–726 (2013).

59. Silverman, R. A. et al. Multicenter study of cigarette smoking among adults with asthma exacerbations in the emergency department, 2011–2012. Respir. Med. 125, 89–91 (2017).

60. McClave, A. K., McKnight-Eily, L. R., Davis, S. P. & Dube, S. R. Smoking Characteristics of Adults With Selected Lifetime Mental Illnesses: Results From the 2007 National Health Interview Survey. Am. J. Public Health 100, 2464–2472 (2010).

61. Prochaska, J. J., Das, S. & Young-Wolff, K. C. Smoking, Mental Illness, and Public Health. Annu. Rev. Public Health 38, 165–185 (2017).

62. Romo-Nava, F. et al. Revisiting the bipolar disorder with migraine phenotype: Clinical features and comorbidity. J. Affect. Disord. 295, 156–162 (2021).

63. Michel, I. M. et al. Examining the Moderating Effect of Sex on the Association Between Asthma and Age of Onset in Bipolar Disorder and Schizophrenia. J. Acad. Consult.-Liaison Psychiatry 66, 482–491 (2025).

64. Wu, Q. et al. Childhood and Parental Asthma, Future Risk of Bipolar Disorder and Schizophrenia Spectrum Disorders: A Population-Based Cohort Study. Schizophr. Bull. 45, 360–368 (2019).

65. Hiyoshi, A. et al. Precursors in adolescence of adult-onset bipolar disorder. J. Affect. Disord. 218, 353–358 (2017).

66. Choi, S. W. et al. PRSet: Pathway-based polygenic risk score analyses and software. PLOS Genet. 19, e1010624 (2023).

67. Kichaev, G. et al. Improved methods for multi-trait fine mapping of pleiotropic risk loci. Bioinformatics 33, 248–255 (2017).

68. Zou, Y., Carbonetto, P., Xie, D., Wang, G. & Stephens, M. Fast and flexible joint fine-mapping of multiple traits via the Sum of Single Effects model. Preprint at 10.1101/2023.04.14.536893 (2023).

69. Chen, W. et al. Improved analyses of GWAS summary statistics by reducing data heterogeneity and errors. Nat. Commun. 12, 7117 (2021).

70. Rahimian, R., Wakid, M., O’Leary, L. A. & Mechawar, N. The emerging tale of microglia in psychiatric disorders. Neurosci. Biobehav. Rev. 131, 1–29 (2021).

71. Tsuo, K. et al. Multi-ancestry meta-analysis of asthma identifies novel associations and highlights the value of increased power and diversity. Cell Genomics 2, 100212 (2022).

72. Shrine, N. et al. Moderate-to-severe asthma in individuals of European ancestry: a genome-wide association study. Lancet Respir. Med. 7, 20–34 (2019).

73. Ferreira, M. A. R. et al. Genetic Architectures of Childhood- and Adult-Onset Asthma Are Partly Distinct. Am. J. Hum. Genet. 104, 665–684 (2019).

74. Adams, M. J. et al. Trans-ancestry genome-wide study of depression identifies 697 associations implicating cell types and pharmacotherapies. Cell 188, 640–652.e9 (2025).

75. Forstner, A. J. et al. Genome-wide association study of panic disorder reveals genetic overlap with neuroticism and depression. Mol. Psychiatry 26, 4179–4190 (2021).

76. Otowa, T. et al. Meta-analysis of genome-wide association studies of anxiety disorders. Mol. Psychiatry 21, 1391–1399 (2016).

77. Demontis, D. et al. Genome-wide analyses of ADHD identify 27 risk loci, refine the genetic architecture and implicate several cognitive domains. Nat. Genet. 55, 198–208 (2023).

78. Autism Spectrum Disorder Working Group of the Psychiatric Genomics Consortium et al. Identification of common genetic risk variants for autism spectrum disorder. Nat. Genet. 51, 431–444 (2019).

79. Mullins, N. et al. Genome-wide association study of more than 40,000 bipolar disorder cases provides new insights into the underlying biology. Nat. Genet. 53, 817–829 (2021).

80. Nievergelt, C. M. et al. Genome-wide association analyses identify 95 risk loci and provide insights into the neurobiology of post-traumatic stress disorder. Nat. Genet. 56, 792–808 (2024).

81. Yu, D. et al. Interrogating the Genetic Determinants of Tourette’s Syndrome and Other Tic Disorders Through Genome-Wide Association Studies. Am. J. Psychiatry 176, 217–227 (2019).

82. Strom, N. I. et al. Genome-Wide Association Study of Obsessive-Compulsive Symptoms including 33,943 individuals from the general population. Mol. Psychiatry 29, 2714–2723 (2024).

83. International Obsessive Compulsive Disorder Foundation Genetics Collaborative (IOCDF-GC) and OCD Collaborative Genetics Association Studies (OCGAS). Revealing the complex genetic architecture of obsessive–compulsive disorder using meta-analysis. Mol. Psychiatry 23, 1181–1188 (2018).

84. Duncan, L. et al. Significant Locus and Metabolic Genetic Correlations Revealed in Genome-Wide Association Study of Anorexia Nervosa. Am. J. Psychiatry 174, 850–858 (2017).

85. Schizophrenia Working Group of the Psychiatric Genomics Consortium et al. Comparative genetic architectures of schizophrenia in East Asian and European populations. Nat. Genet. 51, 1670–1678 (2019).

86. Johnson, E. C. et al. A large-scale genome-wide association study meta-analysis of cannabis use disorder. Lancet Psychiatry 7, 1032–1045 (2020).

87. Julienne, H. et al. JASS: command line and web interface for the joint analysis of GWAS results. NAR Genomics Bioinforma. 2, lqaa003 (2020).

88. Julienne, H., Shi, H., Pasaniuc, B. & Aschard, H. RAISS: robust and accurate imputation from summary statistics. Bioinformatics 35, 4837–4839 (2019).

89. Troubat, L., Fettahoglu, D., Henches, L., Aschard, H. & Julienne, H. Multi-trait GWAS for diverse ancestries: mapping the knowledge gap. BMC Genomics 25, 375 (2024).

90. Giambartolomei, C. et al. Bayesian Test for Colocalisation between Pairs of Genetic Association Studies Using Summary Statistics. PLoS Genet. 10, e1004383 (2014).

91. Ahlmann-Eltze, C. & Huber, W. glmGamPoi: fitting Gamma-Poisson generalized linear models on single cell count data. Bioinformatics 36, 5701–5702 (2021).

